# Evidence of SARS-Cov-2-specific memory B cells six months after vaccination with BNT162b2 mRNA vaccine

**DOI:** 10.1101/2021.07.12.21259864

**Authors:** Annalisa Ciabattini, Gabiria Pastore, Fabio Fiorino, Jacopo Polvere, Simone Lucchesi, Elena Pettini, Stefano Auddino, Ilaria Rancan, Miriam Durante, Michele Miscia, Barbara Rossetti, Massimiliano Fabbiani, Francesca Montagnani, Donata Medaglini

## Abstract

SARS-CoV-2 mRNA vaccines have demonstrated high efficacy and immunogenicity, but limited information is currently available on memory B cells generation and long-term persistence. Here, we investigated Spike-specific memory B cells and humoral responses in 145 subjects, up to six months after the BNT162b2 vaccine (Comirnaty) administration. Spike-specific antibody titers peaked 7 days after the second dose and significant titers and neutralizing activity were still observed after six months, despite a progressive decline over time. Concomitant to antibody reduction, Spike-specific memory B cells, mostly IgG class-switched, increased in blood of vaccinees and persisted six months after vaccination. Following *in vitro* restimulation, circulating memory B cells reactivated and produced Spike-specific antibodies. A high frequency of Spike-specific IgG^+^ plasmablasts, identified by computational analysis 7 days after boost, positively correlated with the generation of IgG^+^ memory B cells at six months.

These data demonstrate that mRNA BNT162b2 vaccine elicits strong B cell immunity with Spike-specific memory B cells that still persist six months after vaccination, playing a crucial role for rapid response to SARS-CoV-2 virus encounter.

**One Sentence Summary:** mRNA BNT162b2 vaccine elicits persistent spike-specific memory B cells crucial for rapid response to SARS-CoV-2 virus encounter

## INTRODUCTION

Severe acute respiratory syndrome coronavirus 2 (SARS-CoV-2), the agent responsible of Coronavirus Disease 2019 (COVID-19), has infected more than 180 million individuals and is currently responsible of almost 4 million of deaths (https://covid19.who.int/). An unprecedented effort has been made in the development of effective vaccines, essential to prevent further morbidity and mortality. Seventeen different vaccines are currently used worldwide following authorization by national regulatory authorities, seven of which have received the approval for emergency use by the WHO regulatory authority (https://extranet.who.int/pqweb/sites/default/files/documents/Status_COVID_VAX_29June2021.pdf). As of 1 July 2021, a total of 2.950.104.812 vaccine doses have been administered worldwide. Two SARS-CoV-2 vaccines based on the novel messenger RNA (mRNA) technology, BNT162b2 (produced by Pfizer-BioNTech, commercially named Comirnaty) and mRNA-1273 (Moderna), have been licensed for emergency use. This is the first time that mRNA vaccines are approved for human use and limited information is available on the profile and persistence of the immune response elicited. Phase 3 trials of these vaccines have shown an efficacy of 94-95% at preventing symptomatic infection after two doses administered 3 to 4 weeks apart (1, 2). Two recent studies of BNT162b2 vaccine effectiveness against SARS-CoV-2 infection and COVID-19 cases in a nationwide mass vaccination setting, demonstrate that the vaccine is effective for a wide range of COVID-19 related outcomes, such as hospitalization, severe illness, and death (3, 4) confirming data of effectiveness reported in the randomized phase III clinical study (NCT04368728) (1).

However, available data on immunity elicited by mRNA vaccination are mostly related to antibody responses *(5–7)* while limited information is available on persistence of memory B cells that are expected to play a crucial role for rapid response to SARS-CoV-2. The human memory B-cell compartment is indeed a pillar for the vaccine efficacy, since it is the basis of protective immunity upon pathogen encounter and leads to the generation of plasma cells capable to produce spike specific antibodies. Therefore, the assessment of memory B cells provides a critical biomarker to profile the long-term persistence of effective responses even beyond the duration of antibody responses.

How long memory B cell responses elicited by mRNA COVID-19 vaccines will persist still remains therefore a critical open question. To answer this question, we have investigated the generation and persistence of peripheral Spike-specific memory B cells and circulating antibodies up to six months after vaccination with BNT162b2 vaccine in subjects naïve to SARS-CoV-2 infection.

## RESULTS

In this study, we profiled the Spike-specific memory B response and the antibody response following administration of the BNT162b2 SARS-CoV-2 mRNA vaccine in Italian health care workers without a laboratory confirmed history of SARS-CoV-2 infection. A total of 145 subjects were enrolled in the study, of whom 47 (32.4%) were male, median age 47±17 and 98 (67.6%) were female, median age 45.1±12.8. Complete characteristics of volunteers are summarized in Supplementary Table 1. According to the schedule followed for the BNT162b2 mRNA vaccine administration, subjects were vaccinated with two doses, 3 weeks apart. Blood samples were collected at the baseline (day 0), and at days 7, 21 (pre-boost), 28 (7 days post-boost), 90 (3 months after the first vaccination dose), and 160-180 (5-6 months after the first vaccination dose) and assessed for Spike-specific antibodies and memory B cells (Fig. 1).

**Fig. 1:**
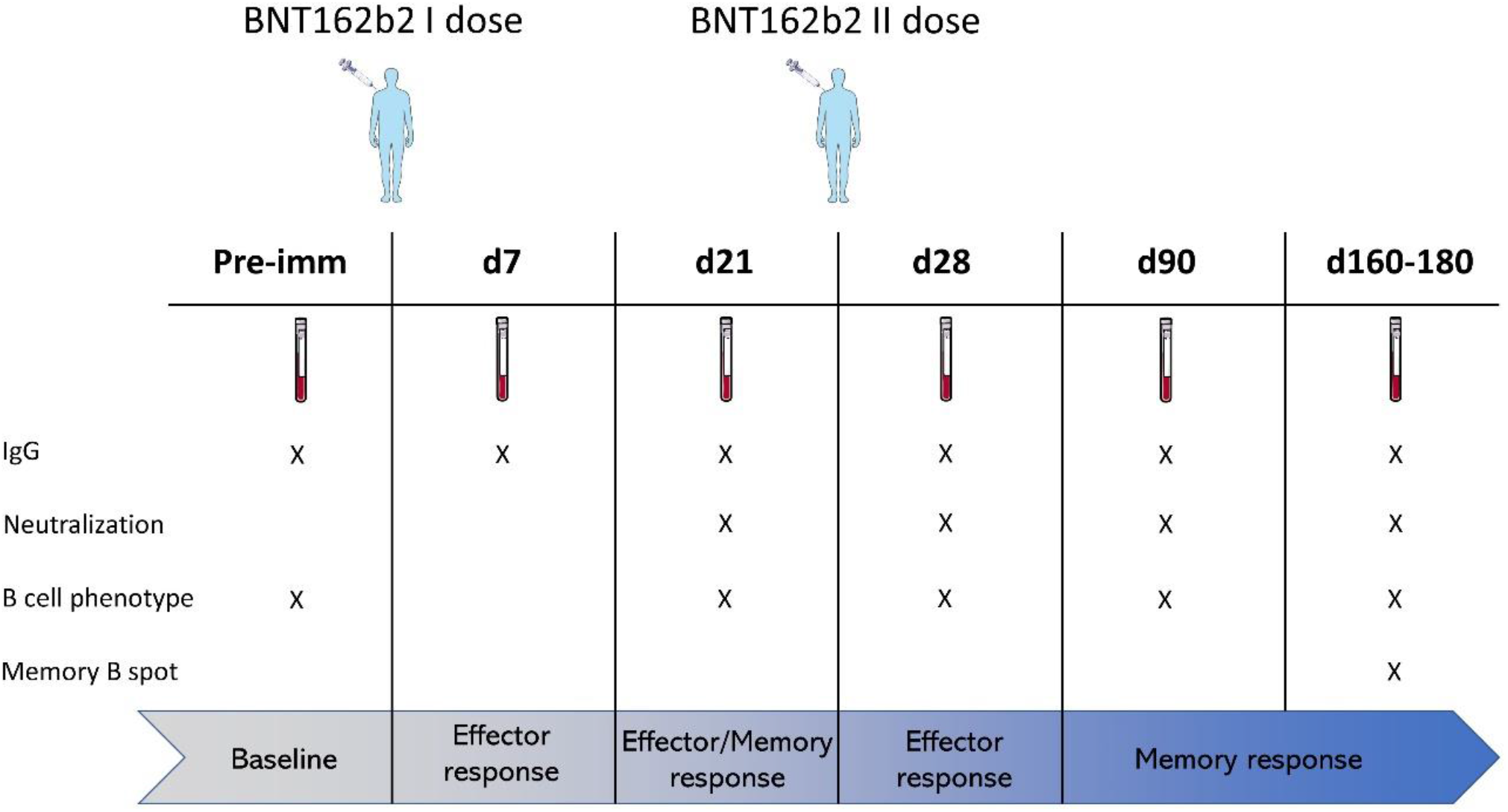
Study design. Healthcare workers (145 subjects) vaccinated with two doses of BNT162b2 mRNA (Pfizer-BioNTech; Comirnaty) vaccine 3 weeks apart were enrolled in the study. Blood samples were collected at the baseline (day 0), 7, 21 (pre-boost), 28 (7 days post-boost), 90 (3 months after the first vaccination dose), and 160-180 (5-6 months after the first vaccination dose) days after vaccination. Plasma and peripheral blood mononuclear cells (PBMCs) were assessed for Spike-specific antibodies and memory B cells, respectively.

### Spike-specific antibody titers and neutralizing activity

Spike-specific IgG levels were assessed in plasma of vaccinated subjects at all the time points. As shown in Figure 2A, a significant increase of Spike-specific IgG levels was observed already after the first dose (21 days), with a geometric mean titer (GMT) of 2869 (95% confidence interval [CI] 2178 to 3778; titers range 160-163840; P≤ 0.001 *versus* baseline and day 7). Antibodies peaked 7 days after the second dose with a GMT of 22120 (95% CI 16319 to 29983; range 320-163840; P≤ 0.001 *versus* baseline). Significant levels were observed also at day 90 (GMT value of 7712; 95% CI 6134 to 9695; range 160-81920; P≤ 0.001 *versus* baseline) and at days 160-180 (GMT value of 3457; 95% CI 2768 to 4317; range 1280-20480; P≤ 0.001 *versus* baseline), despite a progressive significant decline overtime (P≤ 0.05 between days 28 and 90, P≤ 0.001 between days 28 and 160-180; Fig. 2A). No statistically significant difference was observed between days 90 and 160-180.

**Fig. 2:**
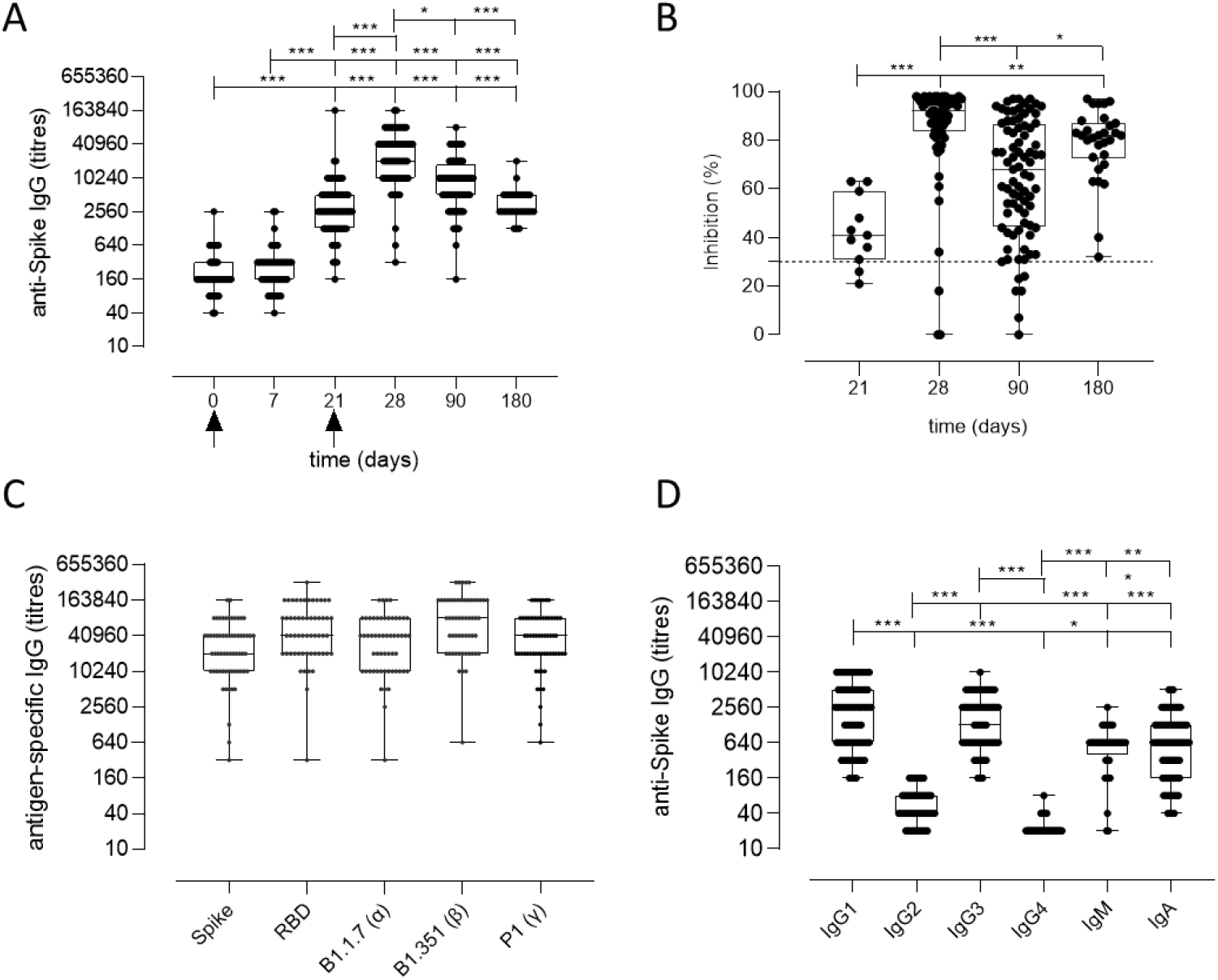
Spike-specific antibody response following BNT162b2 mRNA vaccination. A) Spike-specific IgG analyzed by ELISA in plasma collected 0, 7, 21, 28, 90, 160-180 days after the first dose of the BNT162b2 mRNA vaccine (arrows). Antibody titres are expressed as the reciprocal of the dilution of sample reporting an OD value double respect to the background. Data are shown as box and whiskers diagram showing the minimum and maximum of all the data. B) Surrogate virus neutralization evaluated at days 21, 28, 90 and 160-180 after the first vaccination dose of the BNT162b2 mRNA vaccine. Data are reported as ACE2/RBD inhibition percentage with box and whiskers diagram showing the minimum and maximum of all the data. A threshold (dotted red line) was placed at 30% inhibition percentage to discriminate between neutralization positive and negative samples. C) Titers of IgG anti wild type Spike and RBD, or anti mutated Spike protein (Alpha B.1.1.7, Beta B.1.351, and Gamma P.1) in plasma collected at day 28 (7 days after the second vaccination dose). D) Spike-specific IgG1, IgG2, IgG3, IgG4, IgM and IgA titres in plasma collected at day 28 (7 days after the second vaccination dose). D). Kruskal-Wallis test, followed by Dunn’s post test for multiple comparisons, was used for assessing statistical differences between groups. *P≤ 0.05; **P ≤ 0.01; ***P ≤ 0.001.

Binding of plasma IgG to the receptor-binding domain (RBD) of the Spike protein was also evaluated at day 28 (GMT value of 50791; 95% CI 39232 to 65754) without significant difference compared to the response assessed against the full Spike protein (Fig. 2C).

The analysis of Spike-specific IgG subclasses at day 28 showed a prevalence of IgG1 (GMT of 1780; 95% CI 1280 to 2475; range 160-10240), and IgG3 (GMT of 1402; 95% CI 1066 to 1844; range 160-10240), and a complete absence of IgG2 and IgG4 (GMT of 50 and 22, respectively. Fig. 2D). Low levels of IgA and IgM were detected, with GMT values of 709 (95% CI 521 to 963; range of 40-5120) and 485 (95% CI 346 to 678; range 20-2560) respectively (Fig. 2D).

The age of vaccinated individuals did not significantly impact on the antibody response, even though a higher GMT of 50018 (range 5120-163840) was observed at day 28 in younger individuals (24-30 years) compared to subjects older than 31 years (GMT 33632; range 320-163840; data not shown) in line with other recently reported data *(8, 9)*.

Different SARS-CoV-2 variants, harboring specific mutations in their Spike protein, have emerged during these months. Alpha (B.1.1.7 firstly isolated in United Kingdom), Beta (B.1.351, firstly isolated in South Africa) and Gamma (P.1, firstly isolated in Brazil) variants, were tested against vaccinated plasma collected at day 28 to verify if antibodies generated by the mRNA vaccine were able to bind the mutated Spike protein. As shown in Figure 2C, antibodies efficiently bound the Alpha, Beta and Gamma variants of the Spike protein.

Plasma samples collected at day 21, 28, 90 and 160-180 after vaccination were tested using a surrogate virus neutralization assay (sVN) *(10)* to assess whether antibodies elicited by BNT162b2 mRNA vaccination were capable of blocking RBD-ACE2 interaction thus inhibiting the main entrance way of SARS-CoV2. As shown in Figure 2B, the ACE2/RBD inhibition significantly increased after the second dose (85.48% ±20.50 at day 28 *versus* 42.73 ±14.37 at pre-boost) as well as the percentage of subjects with a positive neutralization value (from 82% after the first dose to 100% after the second dose). From day 28 a decrease of the inhibition percentage value was observed. A moderate positive correlation according to Pearson test (r = 0.6706; P≤ 0.001; Fig S1) was observed between Spike-specific IgG titers and ACE2/RBD inhibition percentage (r = 0.6706; P≤ 0.001; Fig S1).

### Spike-specific memory B cells generation and persistence

The generation of the B cell response upon vaccination is characterized by the induction of subsets of cells with different functionalities and phenotypes. Here, we profiled the Spike-specific B cell response after administration of two doses of the BNT162b2 mRNA vaccine, and we followed the persistence of memory B cells up to 160-180 days after vaccination. According to the different time points, we expected to identify both Spike-specific effector (plasmablasts and plasmacells) and memory cells. SARS-CoV-2 specific B cells were identified among CD19^+^ cells using a fluorescent Spike antigen, and their phenotype was characterized assessing the expression of IgD, CD27, CD38, IgG, IgM, IgA, CD24, CXCR5 and CD11c molecules. Spike-specific cells, assessed before (day 21) and after (day 28, 90 and 160-180) the second dose, were detected within the CD19^+^ cells (hereafter named S^+^ B cells, Fig. 3A) in order to identify all the possible subsets elicited by vaccination. The manual analysis was then compared to the computational one, that allows to profile the B cells in an unbiased way, identifying different cell clusters based on the simultaneous expression of all the surface markers analysed.

**Fig. 3:**
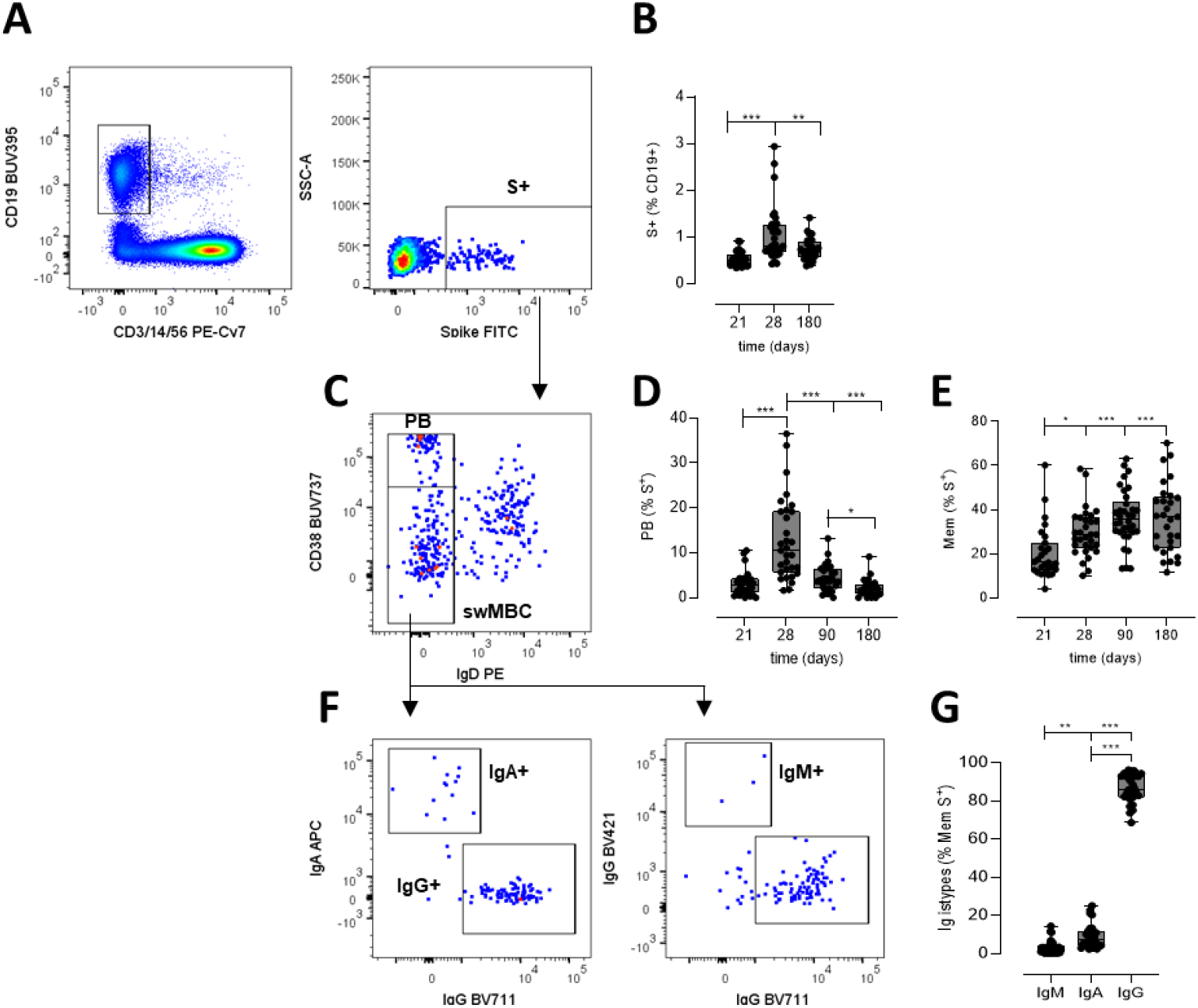
Spike-specific memory B cell response following BNT162b2 mRNA vaccination. Identification of Spike-specific B cells by flow cytometry within PBMCs collected at different time points following vaccine administration. A) Gating strategy for identifying CD19^+^ Spike-specific B cells (named S^+^ B cells) by multiparametric flow cytometry. B) Percentages of S^+^ B cells in each subjects assessed immediately before (day 21) and after (days 28, 90 and 160-180) the second dose. C) Dot plot analysis of CD38 *versus* IgD within S^+^ B cells, for identifying IgD^-^CD38 ^high^ plasmablasts (PB) and Ig-switched memory B cells (swMBC). D-E) Percentages of PB (D) and swMBC (E) in single subjects assessed at day 21, 28, 90 and 160-180. F) Dot plot analysis of IgA, IgG and IgM expression within swMBC. G) Percentages of IgM, IgA and IgG swMBC in single subjects assessed at days 160-180. Dot plots in A), C) and F) are representative from a single subject; values in B), D), E) and G) are reported as box and whiskers diagram showing the minimum and maximum of all of the data; percentages are reported respect to the parent population (in brackets). Statistical difference was assessed by Kruskall Wallis test; *P≤ 0.05; **P ≤ 0.01***; P ≤ 0.001.

S^+^ B cells were significantly increased after the second vaccine dose (P<0.01 *versus* pre-boost) and were maintained significantly higher 6 months after vaccination (P<0.01 *versus* pre-boost, Fig. 3B). The phenotype of the S^+^ B cells changed during the analysis at different time points, in accordance with their effector function. Indeed, the higher frequency of Ig-switched (IgD^-^) CD38^bright^ S^+^ B cells (Fig. 3C), indicative of plasmablasts (14–16), was detected 7 days after the second dose administration (Fig. 3D). These antigen-specific plasmablasts significantly increased compared to the pre-boost (about 13% of S^+^ B cells, *versus* 3% at day 21), and declined overtime (4.3% and 2.5% at 90 and 160-180 days, respectively). On the contrary the switched-memory B cells, identified as IgD^-^ CD38^int/low^ (Fig. 3C), increased after the second dose, starting with a percentage of 16% at day 21 up to 36% at days 90 and 160-180 (Fig. 3E). Within the memory S^+^ B cells detected six months after vaccination, only a low amount expressed IgM (3.2±3.6%) or IgA (8.9±6.3 %), while the majority were switched to IgG (86±7 %) (Fig. 3F).

### Identification of Spike-specific B cell clusters by automated analysis

The computational analysis of multiparametric flow cytometry data is a powerful tool for dissecting all the possible phenotypes present in a sample, in an operator-independent unbiased way *(11)*. To automatically profile the Spike-specific B cells elicited by the mRNA vaccine and determine their modulation at the different time points following vaccine administration, the S^+^ B cells were analyzed employing the FlowSOM clustering algorithm *(12)*. This approach considers the distribution of all surface markers simultaneously, allowing to characterize most of the possible phenotypes in an unbiased way, including the unexpected ones *(13)*.

Different clusters of Spike-specific B cells are visualized in Figure 4A as heatmap showing each FlowSOM metacluster in row and surface markers in column. The percentages of cells positive for each marker inside the cluster is visualized as color-scale from blue (0% of positive cells) to red (100%). The modulation of the different metaclusters at baseline (day 0), before the boosting (day 21), and after the second dose (days 28, 90 and 160-180) is reported in each subject in the heatmap in Figure 4B.

**Fig. 4:**
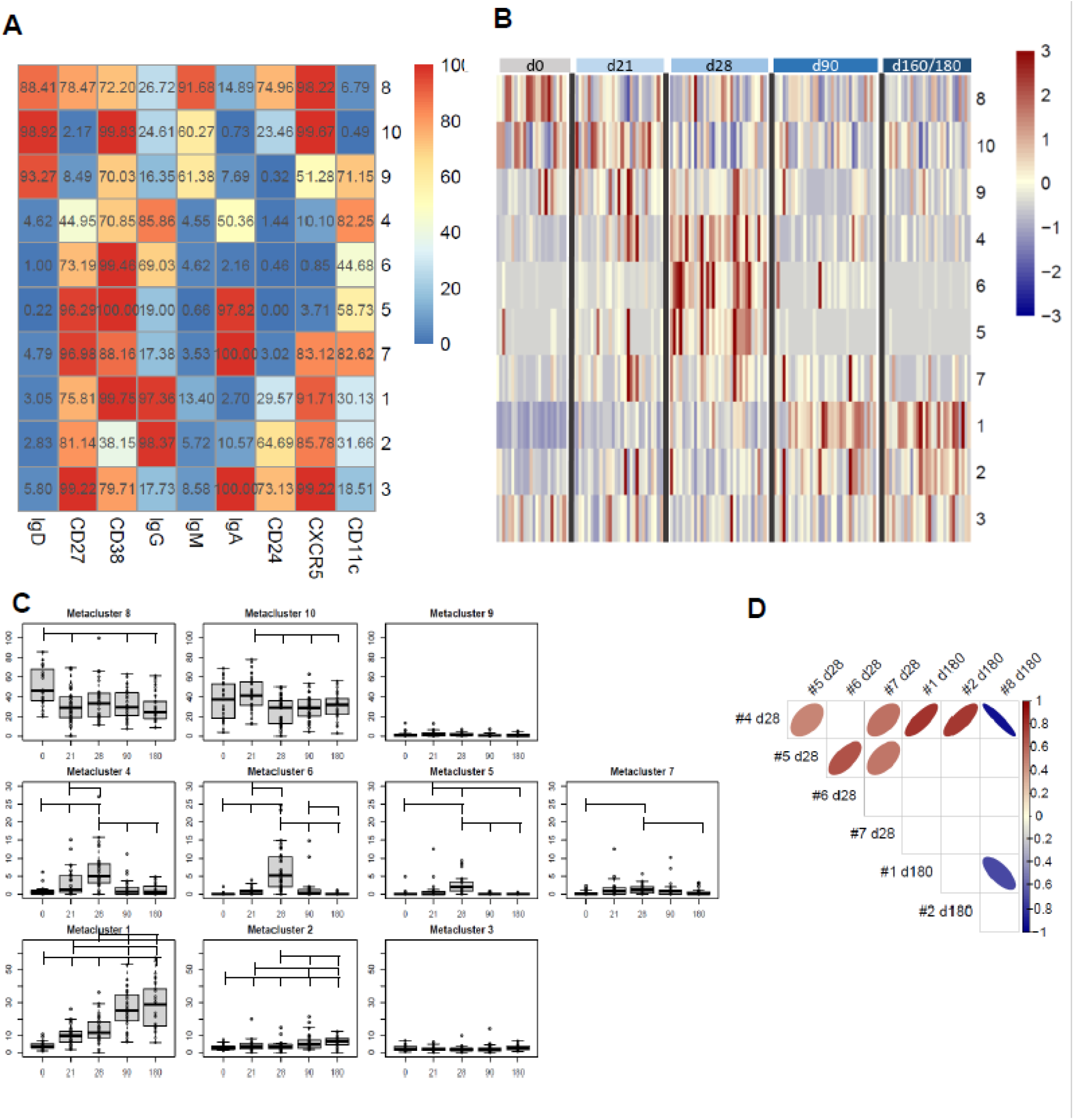
Computational analysis of Spike-specific B cells following BNT162b2 mRNA vaccination. **(A)** Heatmap of metaclusters from FlowSOM analysis of CD19^+^Spike^+^ B cells. Each marker is reported in column, while the different metaclusters are reported in rows. The percentage of cells positive for each marker is reported inside the heatmap boxes, and visualized with a color scale from blue (0%) to red (100%). **(B)** Heatmap reporting the normalized frequency of each sample (columns) for each metacluster (rows) in a color scale from blue (low) to red (high). Columns were grouped by sampling day, as reported above the heatmap. **(C)** Box and whiskers plots showing the frequency of each metacluster at different time points. Values from individual samples were reported as circles. Mann–Whitney test corrected for multiple tests (Benjamini-Hochberg method) was used for assessing statistical differences between different time points; statistical significance was defined as FDR < 10^−2^. **(D)** Correlation analysis between significantly modulated metaclusters at days 28 (4, 5, 6 and 7) and at day 180 (1, 2, and 8). Multiple correlations were visualized as matrix in which ellipses direction and color are proportional to the Spearman’s correlation coefficient between 1 (red) and –1 (blue). Only correlation with FDR<10^−2^ were displayed.

While Spike-specific IgM/IgD double positive subsets (metaclusters 8, 9 and 10) were mostly expressed at baseline or after the first vaccine dose (day 21), Ig-switched memory cells (IgD^-^ CD27^+^; metaclusters 4, 5, 6, 1, 2, 7, 3) significantly increased after the second vaccine dose (Fig 4B-C). Seven days after the vaccine administration, most of the Spike-specific clusters were CD27^+^, CD38^+^ and CD11c^+^ and were negative for CD24 and CXCR5, according to the plasmablast phenotype *(14)*. Some of these plasmablasts were IgA^+^ (metaclusters 5 and 7) and others IgG^+^ (4 and 6). Three and six months after vaccination, the phenotype of most of S^+^ B cells changed into memory cells, as clearly shown by the downregulation of CD11c, the high expression of both CXCR5 and CD24. Even though some of these memory cells were still IgA^+^ (metacluster 3), the majority switched to IgG (clusters 1 and 2) (Fig. 4C).

The automated analysis offers the possibility of analyzing all the phenotypes present within the Spike-positive cells and tracking their modulation according to different time points following the vaccine administration. As clearly shown in Figure 4B, we can observe a time-dependent modulation of the B cell response, that moving from CD27^+^ IgD^+^ IgM^+^ CXCR5^+^ cells at early time points, differentiate into both IgA or IgG effector cells (CD27^+^ IgD^-^ CD38^+^ CXCR5^-^ CD11c^+^) at 7 days after the second dose, and then develop into memory cells (CD27^+^ IgD^-^ CD38^+^ CXCR5^+^ CD24^+^ CD11c^-^), perfectly correlating with the expected phenotypes.

The multiple correlation analysis, performed to assess the relationship between the most significant metaclusters at days 28 and 160-180, showed that the frequency of different plasmablast metaclusters (4, 5, 6 and 7) significantly correlated each other at day 28, and that metaclaster 4 positively correlated with the frequency of IgG^+^ switched memory cells (1 and 2) at day 180, and inversely correlated with the most undifferentiated metacluster 8, at day 180 (Fig. 4D).

This analysis not only corroborates what we observed with the manual gating strategy, but consistently improves the characterization of the B cell subsets elicited by the SARS-CoV-2 vaccination.

### Antibody secretion by reactivated memory B cells

To assess the functionality of circulating S^+^B cells present in blood of vaccinated subjects six months after vaccination, PBMCs were restimulated and the frequency of Spike-specific antibody-secreting cells was determined by ELISpot assay (Fig. 5A). Spike-specific IgG-secreting cells were found in the peripheral blood of 66% of the subjects tested, with a mean frequency of about 1% of the total IgG-secreting cells (Fig. 5B). Circulating spike-specific IgM-secreting B cells were also found, with a percentage of about 20% of the total IgM-secreting cells (Fig. 5C). As shown with the automated analysis, IgM^+^ memory cells were also positive for IgD. These subsets of lymphocytes were excluded from the cells reported in Figure 3G, in which only Ig-switched memory B cells were characterized. As control, PBMCs from the same subjects were restimulated with an unrelated antigen.

**Fig 5:**
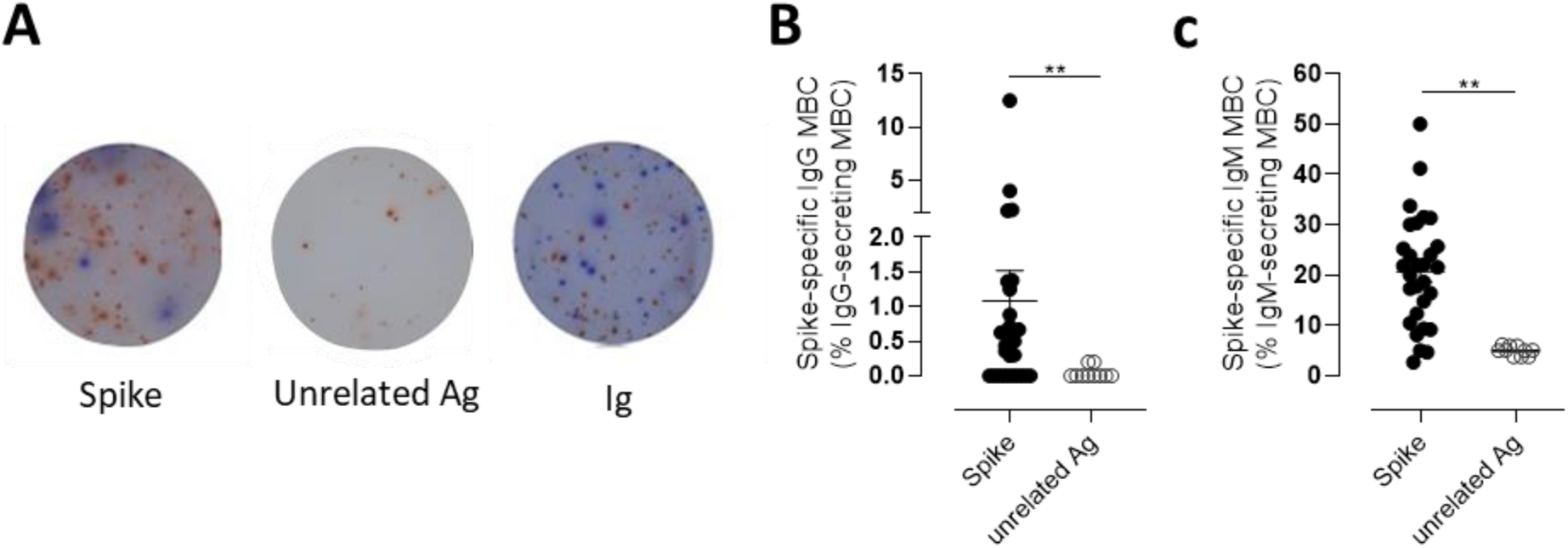
Spike-specific memory B cell response following BNT162b2 mRNA vaccination evaluated by B-cell ELISPOT. **(A)** Representative images of ELISpot wells coated with spike (left), an unrelated antigen (center) or anti-immunoglobulin (Ig, right) and developed in blue and red for IgG and IgM, respectively, after incubation of PBMCs. Cells were collected 160-180 days following the first dose of mRNA BTN162b2 vaccination, and re-stimulated *in vitro* with B-Poly-S for 4 days to induce resting MBC differentiation into antibody-secreting cells. **(B-C)** The frequencies of Spike-specific MBCs secreting IgG (B) or IgM (C) antibodies are reported as percentages of total MBCs producing antibodies of the respective isotype. Bars indicate mean±SEM. Mann–Whitney test, followed by Dunn’s post test for multiple comparisons, was used for assessing statistical difference between Spike-specific and unrelated antigen-specific B cells. **P≤ 0.01.

Taken together, these data well describe the kinetic of the Spike-specific B response elicited by the vaccination with BNT162b2 mRNA vaccine, in which the slow decline of Spike-specific antibody levels overtime is accompanied by the induction of circulating Spike-specific IgM and IgG switched memory B cells, that are still persistent six months after mRNA BTN162b2 vaccination.

## Discussion

In this work we demonstrate that Spike-specific memory B cells, capable of reactivation following antigen encounter, persist in blood of vaccinated subjects six months after the administration of BNT162b2 SARS-CoV-2 mRNA vaccine. Concomitant to antibody reduction, Spike-specific memory B cells, mostly IgG class-switched, increase in blood of vaccinees and persist six months after vaccination. Considering the natural decline of spike-specific circulating antibodies, our results highlight the importance of profiling the antigen-specific memory B cell response, a crucial biomarker of vaccine immunity that could be particularly important to monitor vaccine responsiveness and long-term memory persistence.

While most available data of BNT162b2 vaccine are on antibodies elicited upon the first vaccine dose in healthy or SARS-CoV-2 previously infected subjects or at early time points following the second dose *(18–24)*, our study profiling the antibody response and memory B cells up to six months after vaccination contributes to better understand the BNT162b2 vaccine immunogenicity in SARS-CoV-2 naive subjects.

Through a computational analysis of flow cytometry data, we profiled the Spike-specific B cell response, identifying Spike-specific plasmablasts 7 days after the second vaccine dose, and Ig-switched memory B cells that increased at month 3 and still persisted at month 6 post vaccination. The transient appearance of plasmablasts in blood with a peak at 7 days after BNT162b2 mRNA vaccine administration is in line with what observed with other vaccines, such as attenuated yellow fever strain YF-17D, inactivated influenza vaccine and tetanus vaccine *(14)*.

Most of the Spike-specific memory B cells were IgG^+^, but also IgD/IgM double positive cells were detected. The frequency of Spike-specific IgG^+^ plasmablasts present 7 days post the second vaccine dose positively correlated with the frequency of IgG^+^ memory B cells at day 180, suggesting a predictive value of plasmablast frequency for spike-specific memory B cells generation. Systems biology approaches aimed to identify immunological parameters predictive of long-term responses elicited by vaccination against influenza, have also identified the early induction of plasmablast as a potential biomarker of memory B cells generation *(15)*.

In the context of viral infections it is critical to analyze the persistence of the antibody response and to measure protective antibody titers by functional assays, as well as to assess the ability of circulating memory B cells that can reactivate following antigen encounter. Rapid activation of memory B cells and their differentiation into antibody-secreting PBs is essential for providing antibodies capable of neutralizing the virus *(14)*. When *in vitro* restimulated, circulating memory Spike-specific B cells elicited by BNT162b2 vaccine were capable of reactivation and differentiation into IgG or IgM-secreting cells in 66% of vaccinated subjects.

Parallel to the dissection of the B cellular response, we monitored antibody responses against Spike protein for up to six months after vaccination, observing a peak of IgG 7 days after boost and the persistence of significant levels up to six months, despite a progressive decline overtime. Nevertheless, IgG in plasma collected six months after vaccination still inhibited the *in vitro* binding between RBD and the ACE2 receptor, in all the samples assessed. This surrogate virus neutralization assay has been demonstrated to concord with the gold standard 90% plaque reduction neutralization tests (PRNT_90_) for SARS-CoV-2 antibody detection in human sera *(16)*. IgG1 and IgG3 were the most abundant IgG subclasses produced, but also Spike-specific IgA were released according to the detection of IgA^+^ plasmablasts and memory cells at day 28 and 180, respectively. A similar distribution of IgG subclasses has been previously observed in SARS-CoV-2 infected patients and has been negatively associated with viral load in nasopharyngeal swab *(17)*.

To summarise, six months after vaccination with two doses of BNT162b2 vaccine, subjects without a previous history of SARS-CoV-2 infection, concomitant to antibody reduction develop a consistent and persistent Spike specific IgG-memory B cell response, with cells capable of reactivation following antigen encounter. Spike specific IgG antibodies are still present in blood, with a demonstrated neutralization activity, even though the trend of the antibody curve shows a physiological decline overtime. The induction and the longevity of circulating Spike-specific memory B cells, capable of reactivating into a novel wave of plasmablasts and plasmacells producing Spike-specific antibodies following a secondary antigen encounter, is the critical biomarker indicating the capacity of an effective response to pathogen encounter. These data contribute to provide an answer to the open question on the duration of the memory response to the BNT162b2 vaccine and on the possible need and timing of repeated booster doses of a COVID-19 vaccine.

Finally, this type of analysis could be particularly relevant when applied to fragile patients that, due to immune impairment associated with their primary disease *(25)* and/or age *(26–28)*, are particularly at high risk for severe disease and death related to COVID-19. In these subjects the immune response elicited by vaccination can be affected by disease, age and treatment, therefore the possibility of investigating not only the antibody response but characterizing also the behavior of their memory B cells compartment can be instrumental for defying the vaccination policy most adequate for specific patient category.

In conclusion, these data demonstrate that the mRNA BNT162b2 vaccine elicits robust B cell immunity six months after vaccination, with persistent Spike-specific memory B cells crucial for rapid response to Sars-CoV-2 virus encounter and are particularly important to guide vaccination schedules and policies.

## METHODS

### Study design

Plasma and peripheral blood mononuclear cells (PBMCs) samples were obtained from 145 healthcare workers (HCWs) aged 24-75 years who received two doses of BNT162b2 (Pfizer-BioNTech; Comirnaty) vaccine at 3 weeks apart, enrolled at different time point as reported in Table S1. Exclusion criteria included pregnancy, previous documented SARS-CoV-2 infection and immunocompromising comorbidities (congenital, acquired or drug-related). All participants provided written informed consent before participation to the study. Study participants were recruited at the Infectious and Tropical Diseases Unit, Azienda Ospedaliera Universitaria Senese (Siena, Italy). The study was performed in compliance with all relevant ethical regulations and the protocol was approved by local Ethical Committee for Clinical experimentation (CEASVE), protocol code IMMUNO_COV. Clinical data collection and management were carried out using the software REDCap (Research Electronic Data Capture, Vanderbilt University).

### PBMCs isolation

Venous blood samples were collected in a heparin coated blood tube (BD Vacutainer) at the baseline (day 0), and at days 7, 21 (pre-boost), 28 (7 days post-boost), 90 (3 months after the first vaccination dose), and 160-180 (5-6 months after the first vaccination dose). PBMCs were isolated by density-gradient sedimentation, using Ficoll-Paque (Lymphoprep, Voden Medical Instrument, Meda, Italy). Isolated PBMC were then cryopreserved in cell recovery medium [10% DMSO (Thermo Fisher Scientific) and 90% heat inactivated fetal bovine serum (Sigma Aldrich)] and stored in liquid nitrogen until used. Plasma samples were stored at −80°C.

### ELISA

Maxisorp microtiter plates (Nunc, Denmark) were coated with recombinant wild type or mutated SARS-CoV-2 Spike S1+S2 ECD, or Spike RBD (all from Sino Biological) with 50 μl per well of 1 μg/ml protein solution in PBS (Sigma-Aldrich) overnight at 4°C. Plates were blocked at room temperature (RT) for 1 h with 200 μl of 5% skimmed milk powder, 0.05% Tween 20, 1×PBS. All plasma samples, heated at 56°C for 1 hour to reduce risk from any potential residual virus, were added and titrated in twofold dilution in duplicate in 3% skimmed milk powder, 0.05% Tween 20, 1×PBS (diluent buffer) and incubated 1 h at RT. Anti-human horseradish peroxidase (HRP)-conjugated antibodies for IgG (diluted 1:6000), IgM, IgA (diluted 1:4000), IgG1, IgG2, IgG3, IgG4 (diluted 1:2000; all from Southern Biotechnology) were added in diluent buffer for 1 h at RT. Plates were developed with 3,3’,5,5’-Tetramethylbenzidine (TMB) substrate (Thermo Fisher Scientific) for 10 min at RT, followed by addition of 1M stop solution. Absorbance at 450 nm was measured on Multiskan FC Microplate Photometer (Thermo Fisher Scientific). A WHO international positive control (plasma from vaccinated donor diluted 1:5000; NIBSC 20/268) and negative control (plasma from unvaccinated donor diluted 1:20, NIBSC) were added in duplicate to each plate as internal control for assay reproducibility. Antibody end point titres were expressed as the reciprocal of the sample dilution reporting double the background OD value.

### ACE2/RBD inhibition assay

ACE2/RBD inhibition was tested with a SARS-CoV 2 surrogate virus neutralization test (sVNT) kit (cPass™, Genscript), according to the manufacturer protocol. Briefly, plasma samples, positive and negative controls were diluted 1:10 in dilution buffer, mixed 1:1 with HRP-RBD buffer and incubated for 30 min at 37°C. 100 µl of each mixture were added to each well of a ACE2-coated 96 wells flat-bottom plate and incubated for 15 min at 37°C. The plate was washed four times with wash solution and tapped dry. 100 µl of TMB solution were added to each well and the plate was developed for 15 min at room temperature (20-25°). After that, the reaction was quenched adding 50 µl of the stop solution to each well and the OD at 450 nm was instantly read with Multiskan FC Microplate Photometer (Thermo Fisher Scientific). Results of the ACE2/RBD inhibition assay are expressed as follows: percentage inhibition = (1 -sample OD value/negative control OD value) * 100. Inhibition values ≥30% are regarded as positive results, while values <30% as negative results, as indicated by the manufacturer.

### Multiparametric flow cytometry

Two million PBMCs were incubated with BD human FC block (BD Biosciences) for 10 min at RT. Cells were stained with SARS-CoV-2 Spike S1+S2 ECD-His recombinant biotinylated-protein (2 5µg/ml, Sino Biological) in staining Buffer [Phosphate buffered saline (PBS), 0.5% Bovine Serum Albumin (BSA) and 2 mM EDTA, all from Sigma-Aldrich] for 30 min at 4°C, and then stained with FITC-conjugated Streptavidin for 30 min at 4°C. Cells were washed and stained for 30 min at 4°C with the following antibody-fluorochrome panel: CD3-PECy 7 (clone SK7); CD56-PECy7 (clone B159), CD14-PECy7 (clone M5E2), CD19-BUV395 (clone SJ25C1), IgM-BV421 (clone G20-127), IgD-PE (clone IA6-2), CD11c-BB700 (clone 3.9), CXCR5-BV650 (clone RF8B2), CD27-APC-R700 (clone M-T271), CD24-BV786 (clone ML5), CD38-BUV737 (clone HB7), IgG-BV711 (clone G18-145), CD138-PE-CF594 (clone MI15) (all from Becton Dickinson), IgA-APC (clone IS11-8E10, Miltenyi Biotec). Following surface staining, cells were washed once with PBS and labeled with Live/Dead FSV780 according to the manufacturer instruction (BD Biosciences). Cells were washed once with PBS, resuspended in 100 µL BD fixation solution (BD Biosciences) and incubated at 4°C for 15 min in the dark. All antibodies were titrated for optimal dilution. About 1-2 × 10^6^ cells were acquired and stored for each sample with SO LSRFortessa X20 flow cytometer (BD Biosciences). Data analysis was performed using FlowJo v10 (TreeStar, USA).

### Memory B cell ELISpot

PBMCs were collected from HCWs 160-180 days following the first dose of mRNA BTN162b2 vaccination and evaluated for IgM and IgG production using Human IgM/IgG Double-Color Enzymatic ELISpot assay (CTL Europe GmbH, Bonn, Germany). The protocol was performed according to the manufacturer instruction. PBMCs were cultured in complete RPMI medium at a concentration of 2 × 10^6^ PBMCs/ml in 24-well tissue culture plates and polyclonally stimulated to induce antibody production from resting memory B cells (MBC). Cells were stimulated with Polyclonal B cell Stimulator (B-Poly-S, diluted 1:1000) in CTL-Test B medium for 4 days, at 37°C with 5% CO_2_. After stimulation, cells were harvested, pelleted by centrifugation, and counted using the automated cell counter (Bio-Rad Laboratories, USA). Multiscreen filter 96 well plates were pre-wetted with 70% ethanol and then coated with recombinant wild type SARS-CoV-2 Spike S1+S2 ECD (Sino Biological, 10 μg/ml) for the detection of antigen-specific IgG or with anti-Ig capture antibody for the detection of total IgG and IgM overnight at 4 °C. Coating with an unrelated antigen was included. Coated wells were washed with PBS, and 50 µl of CTL-Test B medium supplemented with 1% L-glutamine (Sigma Aldrich) were added to each well. The amount of 1 × 10^6^, 2 × 10^5^, 4 × 10^4^ and 1 × 10^5^, 2 × 10^4^, 4 × 10^3^ pre-stimulated cells were seeded to evaluate spike-specific and total Ig respectively in a volume of 50 µl/well of CTL-Test B medium. After an overnight incubation at 37°C in the presence of 5% CO_2_, cells were then removed by washing with PBS-0.05% Tween 20, and anti-human IgM/IgG detection solution was added for 3 hrs at RT. Plates were washed, incubated with 80 µl/well of Tertiary Solution (FITC-HRP and Strep-AP, both diluted 1:1000) for 1 hr at room temperature, washed again and Blue and Red Developer Solutions were added, each for 15 min. The reaction was stopped by extensive washing in tap water, and plates were dried in the dark at room temperature. The number of spots was determined by plate scanning and analysis performed with an Immunospot S6 Ultimate Analyzer (CTL Europe GmbH).

### Computational flow cytometry analysis

The B cell population analyzed in our data set was gated as live, singlet, CD3^−^/CD14^-^/CD56^-^ CD19^+^ Spike^+^ cells using FlowJo v10 (TreeStar, USA). The analysis was then carried on R platform (v4.0.3). Flow cytometry standard (FCS) files were exported as uncompensated data in R environment as flowSet object (list of FCS), that was then compensated with FlowCore package 2.0.1 *(29)* and logicle transformed *(30)* using the estimateLogicle function for automatic parameters selection for each fluorescence marker. Clustering analysis was performed following the FlowSOM function pipeline (FlowSOM package v1.20.0). Marker expression was normalized with z-score and grid size was set to 7×7. Similar nodes were merged (metaclustering step) setting number of metaclusters to 10. The Euclidean distance was used in both the FlowSOM clustering and metaclustering. Thresholds to bisect positive and negative cells for each marker expression were automatically set with flowDensity package *(31)*. FlowSOM results were displayed as an heatmap reporting the percentage of positive cells for each marker within the metacluster as preoulsy described *(13)*.

### Statistics

Kruskal-Wallis test, followed by Dunn’s post test for multiple comparisons, was used to assess the statistical differences of ELISA titers and ACE2/RBD inhibition percentages at different time points post vaccination. Pearson test was used to evaluate the correlation between log-transformed ELISA titers and ACE2/RBD inhibition for each time point. Kruskal-Wallis test, followed by Dunn’s post test for multiple comparisons, was used for assessing statistical difference between frequencies of Spike-specific B cells assessed by flow cytometry data. Mann–Whitney test was used for assessing statistical difference between clusters at different time points on data of computational flow cytometry analysis. *P*-values were corrected for multiple tests with Benjamini-Hochberg False Discovery Rate (FDR) method *(32)*. Statistical significance was defined as FDR < 10^−2^. Multiple correlations between metaclusters were performed using Spearman’s correlation with psych package and visualized with corrplot package; p-values were corrected with Benjamini-Hockberg FDR method. Mann–Whitney test, followed by Dunn’s post test for multiple comparisons, was used for assessing statistical difference between Spike-specific and unrelated antigen-specific B cells in ELISPOT data. A P-value ≤ 0.05 was considered significant. Analyses were performed using GraphPad Prism v9 (GraphPad Software, USA)

## Data Availability

All data are available in the main text or the supplementary materials.

## Acknowledgments

We would like to thank all the healthcare workers who participated to the study, Ludovica Soldateschi, Roberta Vanni and Serena Vastola for the technical support and the Infectious and Tropical Diseases Unit Nursing staff who chose to cooperate for blood withdrawal. We thank the Bioengineering Laboratory, University of Siena, for the access to Redcap database; Alberto Balistreri and Massimiliano Marrone for the informatics support.

## Author contributions

Conceptualization: AC, GP, MF, FM, DM

Methodology: AC, GP, FF, MD, FM, DM

Investigation: AC, GP, FF, JP, SL, EP, SA, DM

Resources: GP, IR, MM, BR,

Funding acquisition: DM

Project administration: AC, GP, FM, DM

Supervision: AC, FM, DM

Writing – original draft: AC, GP, FF, DM

Writing – review & editing: all the authors

## Data and materials availability

All data are available in the main text or the supplementary materials.

**Table S1.** Characteristics of the study cohort.

|  | **TOTAL** | **PRE-VACCINE** | **POST-VACCINE**<br>**DOSE 1** | **POST-VACCINE**<br>**DOSE 2** |
| --- | --- | --- | --- | --- |
| N | 145 | 63 (43.5%) | 80 (55.2%) | 102 (70.3%) |
| Age In Years, Mean (SD) | 48.8 (14.3) | 47.7 (14.9) | 41.7 (12.5) | 44.2 (13.7) |
| Sex, N |  |  |  |  |
| Female | 98 | 37 | 55 | 72 |
| Male | 47 | 26 | 25 | 30 |

**Figure S1:**
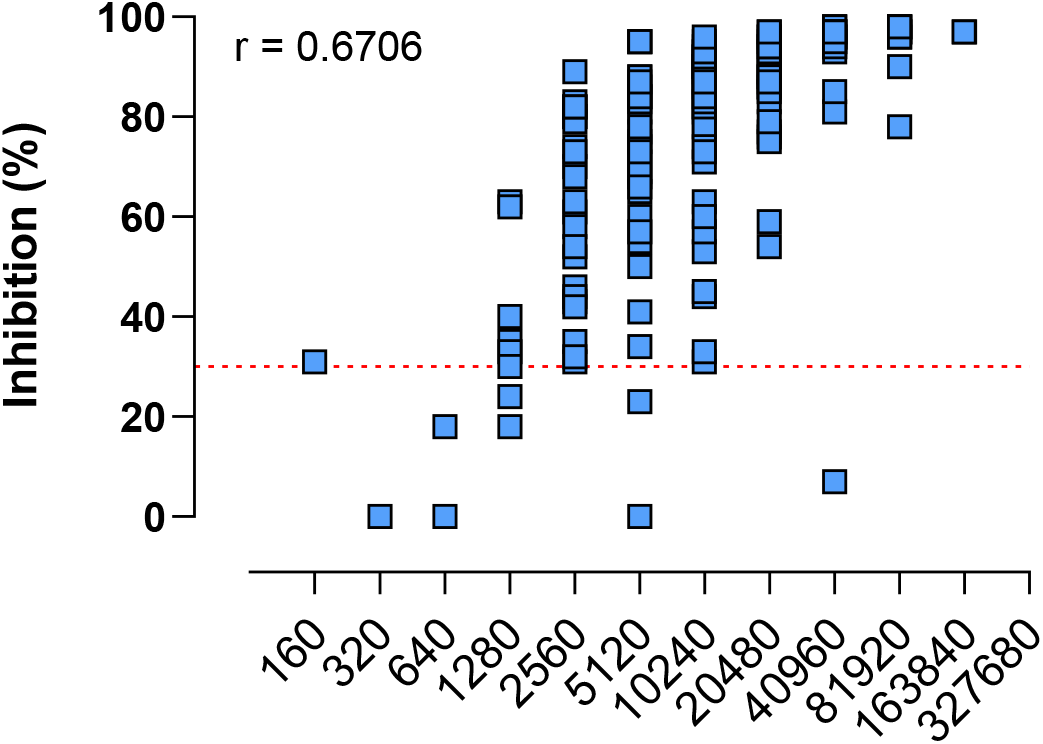
Correlation between IgG ELISA titres and surrogate virus neutralization. Antibody titres are expressed as the reciprocal of the dilution of sample reporting an OD value double respect to the background. The surrogate virus neutralization was expressed as ACE2/RBD inhibition percentage. Pearson correlation test was used to assess the correlation between the two data groups.

